# Sentinel seroprevalence of SARS-CoV-2 in the Gauteng province, South Africa August to October 2020

**DOI:** 10.1101/2021.04.27.21256099

**Authors:** Jaya A George, Siyabonga Khoza, Elizabeth Mayne, Sipho Dlamini, Ngalula Kone, Waasila Jassat, Kamy Chetty, Chad Centner, Taryn Pillay, Mpho R Maphayi, Dineo V Mabuza, Innocent Maposa, Naseem Cassim

**Affiliations:** Department of Chemical Pathology, Faculty of Health Sciences, University of Witwatersrand and National Health Laboratory Service (NHLS), 7 York Road, Parktown, Johannesburg, South Africa; Department of Immunology, Faculty of Health Sciences, University of Witwatersrand and National Health Laboratory Service (NHLS), 7 York Road, Parktown, Johannesburg, South Africa; Division of Public Health Surveillance and Response, National Institute for Communicable Disease of the National Health Laboratory Service, Johannesburg, South Africa; National Health Laboratory Service (NHLS), 7 York Road, Parktown, Johannesburg, South Africa; Division of Medical Microbiology, Groote Schuur Hospital, National Health Laboratory Service and University of the Witwatersrand, Cape Town, South Africa; Division of Epidemiology and Biostatistics, School of Public Health, Faculty of Health Sciences, University of Witwatersrand, 7 York Road, Parktown, Johannesburg, South Africa; Department of Molecular Medicine and Haematology, Faculty of Health Sciences, University of Witwatersrand and National Health Laboratory Service (NHLS), 7 York Road, Parktown, Johannesburg, South Africa

## Abstract

**Background:** Estimates of prevalence of anti-SARS-CoV-2 antibody positivity (seroprevalence) are for tracking the Covid-19 epidemic and are lacking for most African countries.

**Objectives:** To determine the prevalence of antibodies against SARS-CoV2 in a sentinel cohort of patient samples received for routine testing at tertiary laboratories in Johannesburg, South Africa

**Methods:** This sentinel study was conducted using remnant serum samples received at three National Health Laboratory Services laboratories situated in the City of Johannesburg (COJ) district, South Africa. Collection was from 1 August until the 31 October 2020. We extracted accompanying laboratory results for haemoglobin A1c, creatinine, HIV, viral load, and CD4+ T cell count. An anti-SARS -CoV-2 targeting the nucleocapsid (N) protein of the coronavirus with higher affinity for IgM and IgG antibodies was used. We reported crude as well as population weighted and test adjusted seroprevalence. Multivariate logistic regression method was used to determine if age, sex, HIV and diabetic status were associated with increased risk for seropositivity.

**Results:** A total of 6477 samples were analysed; the majority (5290) from the COJ region. After excluding samples with no age or sex stated, the model population weighted and test adjusted seroprevalence for COJ (N=4393) was 27.0 % (95% CI: 25.4-28.6%). Seroprevalence was highest in those aged 45-49 [29.8% (95% CI: 25.5-35.0 %)] and in those from the most densely populated areas of COJ. Risk for seropositivity was highest in those aged 18-49 as well as samples from diabetics (aOR =1.52; 95% CI: 1.13-2.13; p=0.0005) and (aOR=1.36; 95% CI: 1.13-1.63; p=0.001) respectively.

**Conclusion:** Our study conducted during the first wave of the pandemic shows high levels of infection among patients attending public health facilities in Gauteng.

## INTRODUCTION

Severe acute respiratory syndrome coronavirus 2 (SARS-CoV-2) infection is responsible for the global coronavirus disease 2019 (COVID-19) outbreak ^[1]^. The World Health Organisation (WHO) declared COVID-19 a global pandemic on 11 March 2020 ^[2]^. In South Africa, the first case of COVID-19 was reported on 5 March 2020 ^[3, 4]^. To limit the spread of the disease the national government introduced a national lockdown with a five level COVID-19 alert system, starting with alert Level 5 on 27 March 2020. This containment measure delayed the peak of COVID-19 disease^[4]^; however, as restrictions eased, most of South Africa experienced its first peak in July 2020^[5]^.

As of 9 January 2021, South Africa had 1,214,176 laboratory confirmed COVID-19 cases ^[6]^. Confirmation of cases is by amplification and detection of SARS-CoV-2 RNA, in respiratory samples, using real-time reverse transcription polymerase chain reaction (rRT-PCR) assays. PCR testing is mostly offered to both symptomatic persons and contacts (defined as asymptomatic persons who have had close contact with a confirmed COVID-19 case) ^[7]^. This testing strategy precludes the ability to estimate the true burden of disease and importantly cannot detect disease in recovered individuals.

Seroprevalence surveys may provide a better understanding of the epidemiology of COVID-19 than PCR testing. The detection of Anti-SARS CoV-2 antibody testing is able to assess the true number of infections more accurately. The majority of these antibody studies from around the world were conducted early in the epidemic, and were from Europe, Americas and Asia with seroprevalence ranging from <0.1 amongst blood donors in San Francisco to just below 40% from a national seroprevalence survey in India ^[8, 9]^. There is a paucity of similar studies in Africa ^[10, 11]^. To our knowledge, there is one completed South African study, in Cape Town, which examined the seroprevalence in a sentinel survey of public sector patients. They found high seropositivity rates ranging from 31 – 46% in different populations ^[12]^. This study was conducted between July and August of 2020, after the peak of the first wave in Cape Town and utilised convenience sampling. The majority of the patients sampled were primarily attending antenatal or human immunodeficiency virus (HIV) clinical care centres and were therefore overwhelmingly female and adult ^[12]^

South Africa, the African country most severely affected on the continent, has a heavy burden of communicable diseases (especially HIV and tuberculosis) as well as non-communicable diseases (NCD) such as diabetes (DM), hypertension, cancers, and chronic kidney disease ^[13]^. These may all predispose to severe COVID-19 disease ^[14, 15]^.

The aim of this sentinel cross-sectional survey was to estimate the seroprevalence of anti-SARS-CoV-2 antibodies in patients visiting public healthcare centres in the Gauteng province.

## METHODS

### Ethics approval

The study was approved by the University of the Witwatersrand Human Ethics Committee (certificate number M2008105).

### Study design

This was a cross-sectional sentinel study conducted using remnant serum samples received at three National Health Laboratory Services (NHLS) laboratories situated in the City of Johannesburg metropolitan area, South Africa. These laboratories receive samples from surrounding public sector Primary Health Care (PHC) facilities and hospitals.

### Samples selection

We randomly selected remnant samples received at the Chemistry departments from Charlotte Maxeke Johannesburg Academic (CMJAH), Chris Hani Baragwanath Academic (CHBH) and Helen Joseph (HJH) hospitals for analysis of routine chemistry tests. Samples of adequate volume, from hospital outpatient departments and PHC facilities, received from August to October 2020 were selected. Accompanying laboratory results for RT-PCR for SARS CoV-2, haemoglobin A1c (HbA1c), creatinine, HIV, viral load, and CD4+ T cell count were included. These were linked to demographic information extracted from the Laboratory Information System (LIS). Specimens from patients were tested only once.

### Laboratory methods

Anti-SARS CoV-2 antibody testing was performed on the Roche Cobas™ e604 analyser utilising the Roche Elecsys™ SARS-CoV2 chemiluminescent assay (both Roche Diagnostics, Mannheim, Germany). The assay measures total antibodies targeting the nucleocapsid (N) protein of the coronavirus with higher affinity for IgM and IgG antibodies. Prior to this survey, the assay was extensively validated utilising a well-characterised selection of positive and negative samples, using local RT-PCR positive symptomatic individuals and stored negative sera taken prior to December 2019 (unpublished data, Grove et al 2020).

### Data preparation

Age categories reported in the 2020 census population estimates were used as follows: (i) 0-4, (ii) 5-9, (iii) 10-14, (iv) 15-19, (v) 20-24, (vi) 25-29, (vii) 30-34, (viii) 35-39, (ix) 40-44, (x) 45-49, (xi) 50-54, (xii) 55-59, (xiii) 60-64, (xiv) 65-69, (xv) 70-74, (xvi) 75-79 and (xvii) 80+. Data without an age or gender value were classified as unknown. Patients were identified as HIV-infected if either an HIV viral load or CD4+ T cell count was recorded or if samples came from an antiretroviral clinic or HIV ward. Patients were classified as diabetic if the sample came from a diabetic clinic and/or the HbA1c ≥6.5%. We divided HIV infected patients by CD4+ T cell count as below the normal reference range (≤500 cells/µl) or within the normal reference range (>500 cells/µl). A CD4+ T cell count above 500 cells/µl CD4+T cell was classified as immune reconstitution. The HIV viral load data was classified as <1000 copies/ml (virological suppression) and ≥1000 copies/ml (detectable viraemia). Based on whether the health facility description included the word ‘Hospital’, collected data was classified as either ‘Hospital’ or ‘Other’ (facility type). All data received as text were converted (coded) to numbers.

### Statistical analysis

Data were captured on Microsoft Excel spreadsheet and analysis was performed in Stata statistical software (Stata Corp, College Station, TX, USA). We reported the number of patients by sex, age category, month and geographic area (health district or regions for City of Johannesburg). We reported the number of patients with a positive SARS-CoV-2 nucleocapsid (N) protein IgG result to determine the crude seroprevalence with the 95% confidence interval (CI) indicated. We reported the same data by facility and ward type (See Figure 1). Furthermore, to improve precision and accuracy of our estimates given a non-random sample, we used a Bayesian logistic multilevel regression model with post stratification weighting (MRP) that included sex as a fixed effect, and age and region as random effects. The Multilevel regression and post stratification model assume heterogeneity in prevalence of seropositivity in the different regions of COJ. The analysis uses the sex-age distribution in each of the COJ regions as weights. After estimating population weighted prevalence, we went further to adjust for the assay uncertainty using the multilevel regression and post stratification model assuming the observed prevalence is a function of assay sensitivity and specificity (more details in supplementary). In seropositivity prevalence analysis, the sensitivity and specificity of the assay are also known to be a source of variation and uncertainty in the estimates. Rjags package (R Foundation for Statistical Computing, Vienna, Austria) was used to fit the MRP models and estimates were reported with associated credible intervals. Jags uses Markov Chain Monte Carlo (MCMC) to generate a sequence of dependent samples from the posterior distribution of the parameters. We reported the population-weighted, and test performance–adjusted SARS-CoV-2 nucleocapsid (N) protein IgG seroprevalence. The model used South Africa Census 2011 population estimates for the City of Johannesburg for administration regions A to G by sex and age category.

**Figure 1:**
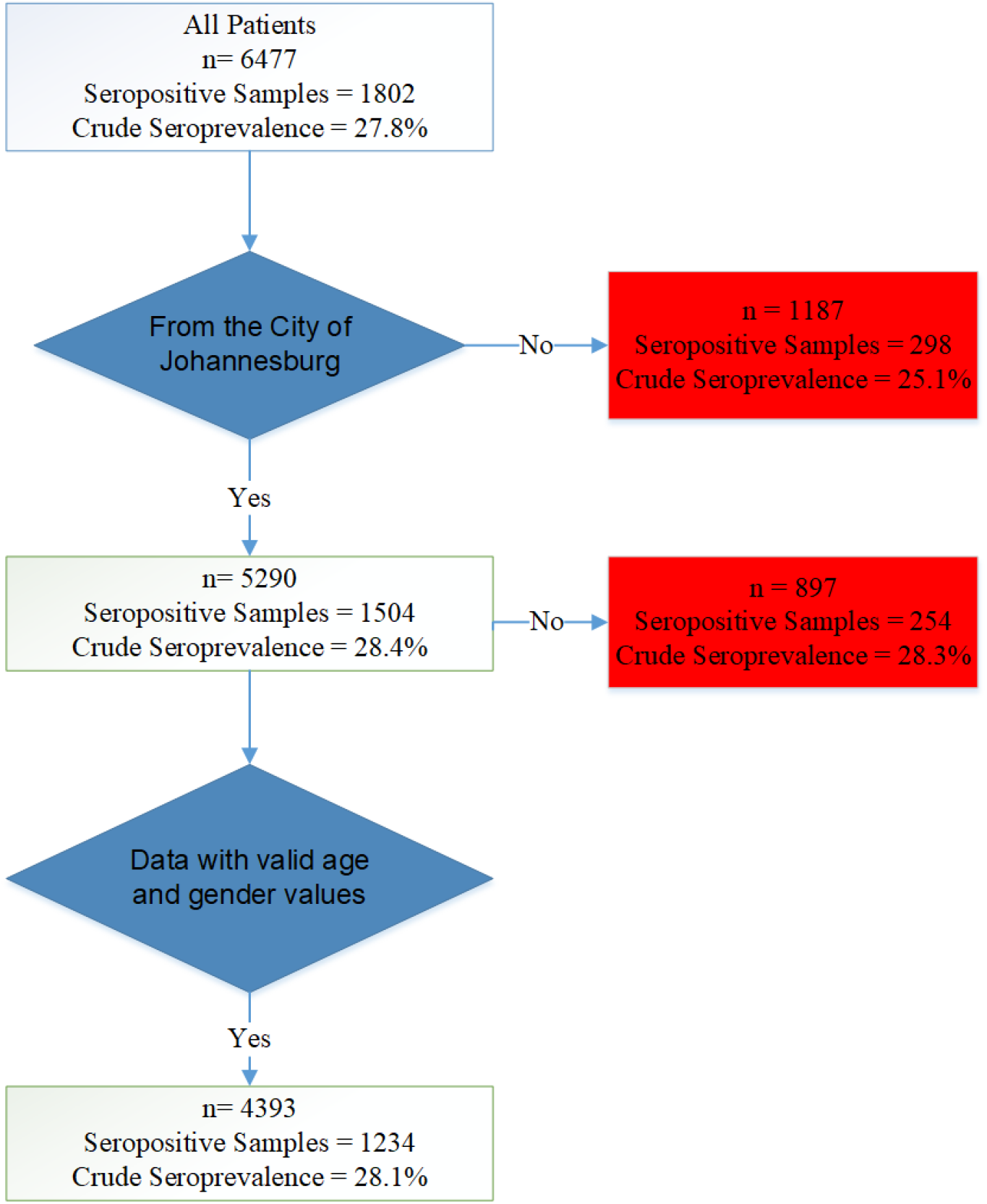
Flow diagram for data analysis indicating how data was prepared.

We compared HbA1c, viral load and CD4 counts between seropositive and seronegative samples. The Wilcoxon rank sum test was used to assess whether the difference in median values was significant. Multivariable logistic regression method was used to determine if age, gender, HIV and diabetic status were associated with increased risk for seropositivity. Age was categorised as <18, 18-49 and >49 years. We reported sex as male and female.

COVID-19 hospitalisations were extracted from DATCOV, a hospital surveillance system. The DATCOV system was used by hospitals for data elements related to admissions and in-hospital deaths. For the August to October 2020 period, we received the numbers of admissions and in-hospital deaths for the City of Johannesburg.

## RESULTS

Supplement Table S1 shows the patient characteristics and crude seroprevalence. A total of 6477 samples was analysed, of which the majority were collected in September 2020 [3347/6477(53.68%)]. Five thousand two hundred and ninety [5290 (81.67%)] of all samples were from the City of Johannesburg District. There were 3234 (49.93%) samples from females. 201 (3.10%) of all samples were from children 4 years or less, and 1091 patients did not have any age stated on the laboratory request form. Crude seroprevalence was highest in August and lowest in September. While, slightly more samples were from patients attending hospital than PHC facilities, the crude seroprevalence was remarkably similar for both (27.8%; 95% CI: 26.3-29.3) versus (27.8%; 95%CI: 26.3-29.5%) respectively.

The crude seroprevalence in HIV positive cases was 26.9% (95% CI: 25.2-28.7%); in diabetics we reported a crude seroprevalence of 32.3% (95%CI: 29.1-35.7), 299 were from women attending ante-natal clinics and the crude seroprevalence was 30.8% (95% CI: 25.6-36.3%). (Supplement Table S2)

To calculate the age and test adjusted seroprevalence for the COJ, we excluded all samples for which there was no age or gender, giving a final number of 4393 samples (Figure 1).

Table 1 shows the adjusted seroprevalence. Adjusted seroprevalence was higher in males, (30.1 %; 95% CI: 26.5-33.2%) than in females (23.5%; 95% CI: 19.4-26.7%). Seropositivity was greater than 20% for all age groups. Seroprevalence was highest in those aged between 45-49 years and was lowest in those 4 years and less. By sub-district, seropositivity was highest in administrative regions E and F which represent the high-density areas of Alexandra, Wynberg, Sandton, Orange Grove, Houghton, Inner City and Johannesburg South (Figure 2).

**Table 1:**
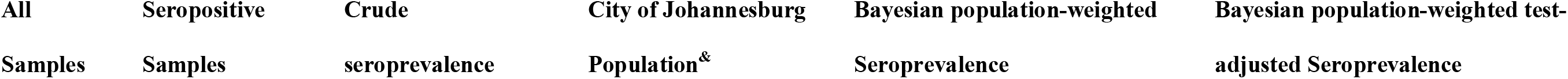
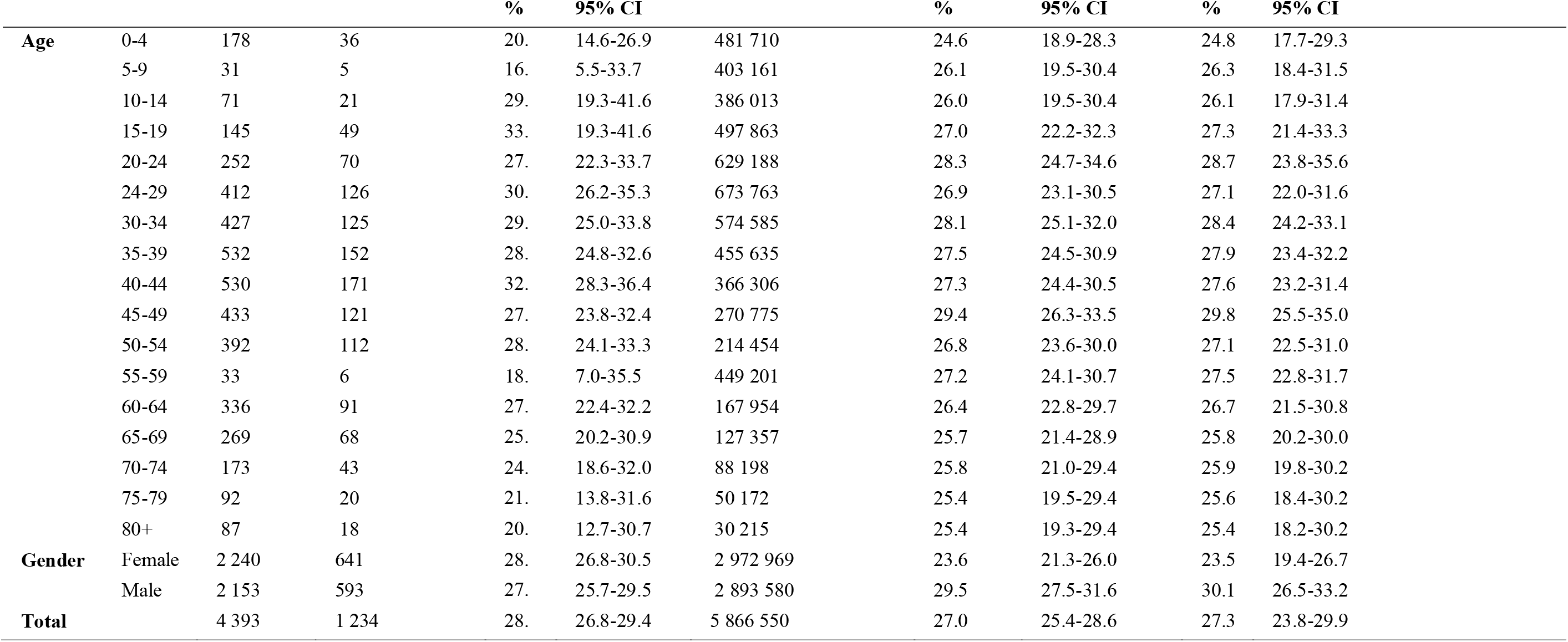
Crude, population-weighted, and test performance–adjusted SARS-CoV-2 nucleocapsid (N) protein IgG seroprevalence for the City of Johannesburg

**Figure 2:**
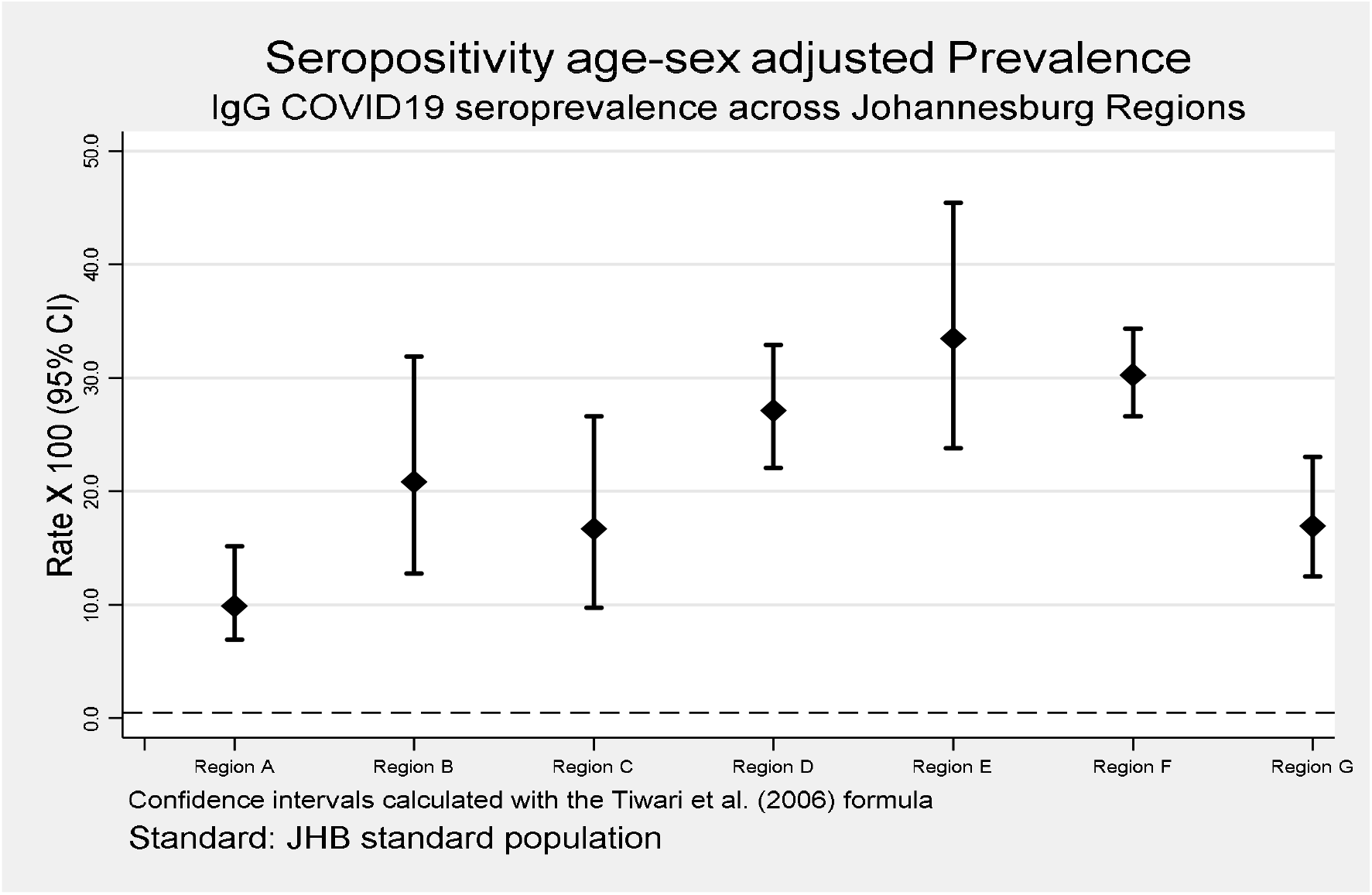
The SARS-CoV-2 nucleocapsid (N) protein IgG Bayesian population-weighted test-adjusted seroprevalence. Legend: Suburbs in the City of Johannesburg that are arranged into the following administrative regions: (i) Region A: Midrand and Northern suburbs (ii) Region B: (iii) Region C: Western suburbs and Roodepoort Fisherville, (iv) Region D: Doornkop, and Soweto (v) Region E: Alexandra, and Sandton (vi) Region F: Inner City and Johannesburg South and (vii) Region G: Orange Farm, and Lenasia

### Trend analysis

Deaths were assessed as a percentage of hospital admissions and these dropped slightly from 21.16% to 14.05% over the study period. At the same time, the crude seroprevalence for the COJ dropped slightly from 31.3% to 26.7%. (Table 2).

**Table 2:** Trend analysis of hospital admissions and seroprevalence for City of Johannesburg

**Johannesburg**
| **Month** | **Hospital admissions** | **Deaths** | **Crude** |
| --- | --- | --- | --- |
| August 2020 | 1049 | 222 (21.16) | 419 /1340(31.1) |
| September 2020 | 415 | 69 (16.62) | 800 /2883(27.7) |
| October 2020 | 256 | 36 (14.05) | 285 / 1067 (26.7) |

Legend: 5290 samples were from COJ. Crude seroprevalence is reported for COJ here along with hospital admissions and deaths for COJ only

### Logistic regression

We carried out multivariable logistic regression to determine if age, HIV status or diabetes increased risk for seropositivity (Figure 3). The risk of seropositivity was similar for males and females (aOR= 0.91; 95% CI: 0.81-1.04; p=0.19). Patients aged 18 to 49 were significantly more likely to be Covid-19 seropositive than the <18 age group, with an odds ratio of 1.52 (95% CI: 1.13-2.13, p=0.67). Diabetics had an increased risk for seropositivity (aOR=1.36; 95% CI: 1.13-1.63, p=0.001) compared to those who were not diagnosed with diabetes.

**Figure 3:**
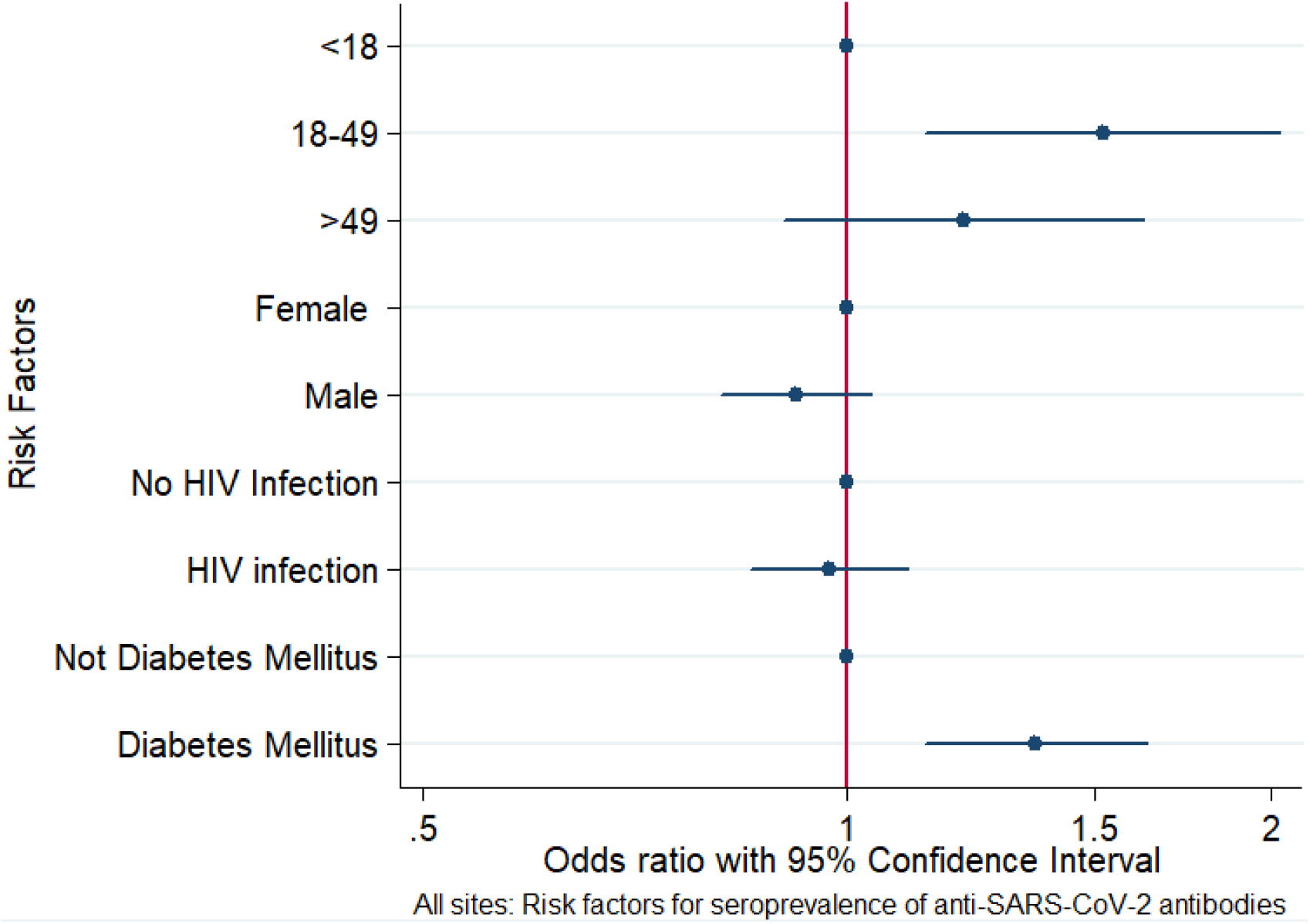
Forest plot reporting the Odds ratio for a positive SARS-CoV-2 nucleocapsid (N) protein IgG test, for various risk factors, between August and October 2020 in the Gauteng province, South Africa.

Finally, we compared HbA1c levels, CD4+ T cell counts and viral load in seropositive and seronegative samples. (Supplement Table S3). We showed that HbA1c was significantly higher in those patients who were seropositive for COVID-19 compared to those who tested negative (8.2%; 95% CI: 6.8-11.0%vs. 7.5% 95% CI: 6.2-9.4%; p=0.02)). CD4+ T cell counts were higher in seropositive patients (625; IQR: 396-653 vs. (576; IQR: 302-641; p=0.0007). Viral load was lower also in seropositive patients, but this was not significant.

## DISCUSSION

We describe a sentinel convenience seroprevalence survey which was undertaken in the Johannesburg metropolitan area in Gauteng, South Africa. This survey was designed to complement other sentinel surveys, most notably the study undertaken in the Western Cape which sampled primarily patients attending routine antenatal and HIV clinical care ^[5]^. Purposive sampling was conducted to ensure that male patients and children were included Overall, the mean age and test adjusted seroprevalence for SARS CoV2 was 27.8% when averaged across all participants studied, with slightly higher rates for males and peak seropositivity in individuals aged between 18 and 49. This was lower than that reported in the Western Cape seroprevalence survey which was 40%. This may reflect the maturity of the Western Cape epidemic which peaked earlier than that in Gauteng. In addition, the higher numbers of international tourists accessing Cape Town in December 2019 and January 2020 may have resulted in earlier and more widespread seeding than suspected.

This survey was conducted from August to October 2020. At this time, PCR confirmed cases of SARS-CoV2 infection were declining with the first wave peak occurring in July 2020^[5]^. The timing of this study is important since detectable serology responses develop at around day 7 and are maximal for IgG after day 14 post-symptom onset ^[16]^. Assuming that the presence of IgG antibodies in this study is at least in the short-term protective, the vast majority of those attending hospitals were still immunologically naïve. The seroprevalence is expected to increase with subsequent waves and a recent study of blood donors in South Africa carried out in 2021 showed a seroprevalence of 60% amongst Black African donors, with varying seroprevalence across provinces ^[17]^. Importantly, our study predates the widespread emergence of the novel 501Y.V2 variant which in early studies is associated with higher transmissibility^[18]^.

Seropositivity in children was not hugely different to that found in adults. As of January 2021, confirmed COVID-19 cases in those under 19 were almost six times lower than that of adults, and admissions were even lower ^[19]^. There are limited data on the seroprevalence of COVID-19 antibodies in children. A study from Bavaria reported similar findings to ours in that antibody positivity in children was higher than actual reported cases of COVID-19 ^[20]^. This has important public health implications for the debates around guidelines for the reopening of schools.

The age and sex adjusted seroprevalence was highest in the more densely populated areas of Johannesburg Inner City, Soweto and surrounds, and Alexandra and surrounds. The overcrowding, often poor sanitation and the impracticability of social distancing, handwashing and mask use are conducive for spread of infection. It has been shown that COVID-19 caseload per million population and per square kilometre are systematically higher in high density areas compared to lower density areas.^[21]^. A limitation of this study is that we do not have any data from private laboratories which serve a different, wealthier population.

Patient comorbidities were considered as an individual predictor of antibody production. In HIV infected individuals (PWH) where data were available, seropositivity correlated significantly with improved immune reconstitution. This is suggestive that HIV-infected individuals are able to produce an antibody response and may be reassuring that PWH who are appropriately controlled will be appropriate vaccine candidates.

Our findings of increased risk for COVID-19 infection in diabetics as evidenced by the presence of antibodies is in keeping with the literature ^[22]^. Furthermore, diabetics are at increased risk of severe disease for multiple reasons, including a compromised innate immunity and the underlying inflammatory state that predisposes to cytokine storm.

COVID-19 infection can also lead to worsening glycaemic control. In South Africa only about 60% of all diabetics are diagnosed and less than 10% are well controlled ^[23]^. We do not have data on the presence of co-morbidities in hospitalized patients. However, in view of the interaction between infectious diseases such as tuberculosis, HIV and now COVID-19 it is imperative that increased attention be paid to the detection and management of non-communicable diseases in South Africa.

A limitation of the study was that it utilised the Roche Elecsys™ kit, which detects antibodies produced against the nucleocapsid. These antibodies do not neutralise the virus and the degree of seroprotection cannot be confirmed. Although recent studies have suggested that nucleocapsid antibodies correlate with levels of neutralising antibodies, this represents an important caveat of this study ^[24]^. The detection of past infection in these individuals however, will be important in assessing the effectiveness of public health and non-pharmacological control methods in South African patients and will assist in informing the vaccine rollout. In addition, it may provide information regarding the potential implications of new variant mutations in South Africa, which may have increased infectivity potential. Furthermore, we used remnant samples from patients attending hospitals or clinics, and therefore this may be an overestimate of seropositivity.

## CONCLUSIONS

Our estimates of seroprevalence, carried out during the first wave of the pandemic are slightly lower than a similar sentinel survey from the Western Cape, showed that almost one in three people from the COJ were infected during the first wave of the COVID-19 pandemic. Combined with household surveys, these results can be used to determine the infection fatality risk, which is a measure of the severity of infection. This is crucial for weighing the risks and benefits of future strategies for the next anticipated waves of infections. Our findings of high seroprevalence in children, high density areas and diabetics have important public health implications. The high seroprevalence in PWH portends well for vaccination in this group of people.

## Data Availability

Data available subject to approval by instututional ethics committee

## ACKNOWLEDGEMENTS

The authors acknowledge the laboratories that provided the remnant samples for our study. We also thank the laboratory staff for conducting the testing and the Division of Public Health Surveillance and Response at the National Institute for Communicable Disease for providing aggregate PCR and DATCOV data.

## SUPPLEMENTARY TABLES

**Table S1:** Patient characteristics and crude seroprevalence

| Variable | Total n= (%) | Positive n= (%) | Crude seroprevalence |  |
| --- | --- | --- | --- | --- |
|  |  |  | % | 95% CI |
| Sex |  |  |  |  |
| Female | 3234 (49.93) | 952 (52.83) | 29.44 | 27.9-31.0 |
| Male | 3 151 (48.61) | 824 (45.73) | 26.15 | 24.6-27.7 |
| Unknown | 92 (1.42) | 26 (1.44) | 28.26 | 19.4-38.6 |
| Age |  |  |  |  |
| 0-4 | 201 (3.10) | 39 (2.16) | 19.4 | 14.2-25.6 |
| 5-9 | 35 (0.54) | 8 (0.44) | 22.9 | 10.4-40.1 |
| 10-14 | 33 (0.51) | 6 (0.33) | 18.2 | 7.0-35.5 |
| 15-19 | 85 (1.31) | 25 (1.39) | 29.4 | 20.0-40.3 |
| 20-24 | 169 (2.61) | 56 (3.11) | 33.1 | 26.1-40.8 |
| 25-29 | 297 (4.59) | 82 (4.55) | 27.6 | 22.6-33.1 |
| 30-34 | 460 (7.10) | 142 (7.88) | 30.9 | 26.7-35.3 |
| 35-39 | 504 (7.78) | 149 (8.27) | 29.6 | 25.6-33.8 |
| 40-44 | 662 (10.22) | 184 (10.21) | 27.8 | 24.4-31.4 |
| 45-49 | 661 (10.21) | 202 (11.21) | 30.6 | 27.1-34.2 |
| 50-54 | 555 (8.57) | 151 (8.38) | 27.2 | 23.5-31.1 |
| 55-59 | 511 (7.89) | 138 (7.66) | 27.0 | 23.2-31.1 |
| 60-64 | 420 (6.48) | 118 (6.55) | 28.1 | 19.5-28.9 |
| 65-69 | 342 (5.28) | 82 (4.55) | 24.0 | 19.5-28.9 |
| 70-74 | 230 (3.55) | 51 (2.83) | 22.2 | 17.0-28.1 |
| 75-79 | 112 (1.73) | 27 (1.50) | 24.1 | 16.5-33.1 |
| 80+ | 109 (1.68) | 25 (1.39) | 22.9 | 15.4-32.0 |
| Unknown | 1 091 (16.84) | 317 (17.59) | 29.1 | 26.4-31.8 |
| Months of collection |  |  |  |  |
| August | 1 460 (22.54) | 436 (24.20) | 29.9 | 27.5-32.3 |
| September | 3 347 (53.68) | 897 (49.78) | 26.8 | 25.3-28.3 |
| October | 1 664 (25.69) | 469 (26.03) | 28.1 | 25.9-30.3 |
| Area |  |  |  |  |
| City of Johannesburg | 5 290 (81.67) | 1 504 (83.46) | 28.4 | 27.2-29.7 |
| Others | 1 187 (18.33) | 298 (16.54) | 25.1 | 22.7-27.7 |
| Total | 6 477 (100.00) | 1 802 (100.00) | 27.8% | 26.7-28.9 |

**Table S2:** Crude SARS-CoV-2 nucleocapsid (N) protein IgG crude seroprevalence reported for facility type and disease.

| Category | Patients N (%) | Positive N (%, CI) |
| --- | --- | --- |
| **Facility Type** |  |  |
| Hospital | 3 431 (53.0) | 954 ((27.8)(26.3-29.3) |
| Other | 3 046 (47.0) | 848 ((27.8)(26.3-29.5) |
| **Ward Type** |  |  |
| Ante-natal clinic | 299 (4.6) | 92 ((30.8)(25.6-36.3) |
| Cancer | 496 (7.7) | 131 ((26.4)(22.6-30.5) |
| Emergency | 415 (6.4) | 117 ((28.2)(23.9-32.8) |
| HIV positive<sup>&</sup> | 2489 (38.4) | 670 ((26.9)(25.2-28.7) |
| Intensive Care Unit | 228 (3.5) | 63 ((27.6)(21.9-33.9) |
| Diabetes<sup>&</sup> | 802 (12.4) | 259 ((32.3)(29.1-35.7) |

<sup>&</sup>Used both the ward type and laboratory results to assign status
Legend: We reported data for wards or clinics for which we had > 200 samples.

**Table S3:** Median and interquartile range for CD4, HIV viral load and HbA1c testing for SARS-CoV-2 nucleocapsid (N) protein IgG crude positive and negative results for August to October 2020 in the Gauteng province, South Africa.

| Test | Positive | Negative | p-value* |
| --- | --- | --- | --- |
| Median CD4 (cells/μl) | 625<br>(396,653) | 576<br>(302,641) | 0.0007 |
| Median HIV Viral Load (copies/ml) | 19<br>(19,27) | 19<br>(19,35) | 0.8125 |
| Hba1c | 8.2<br>(6.8,11) | 7.5<br>(6.2,9.4) | 0.0216 |
| *Wilcoxon rank sum test |  |  |  |

## Statistical analysis supplement

The study reports crude seropositivity prevalence for COVID-19 infection for Gauteng province and then focuses more specifically on Johannesburg district because the bulk of sample were from this district. Since the data was collected non-randomly, using sentinel surveillance, we adjusted the prevalence by using a model-based population weighted framework.

The multilevel logistic regression with poststratification, generally referred to as Multilevel Regression and Poststratification (MRP) was used (Gelman et al, 2016). The model uses age-sex distributions in each region as weights. The population weights were in 238 age-sex-region strata (17 age categories, 2 sex levels and 7 regions). The sentinel survey used samples from all population age groups. The 2011 Johannesburg population from census data were used for the poststratification weights. The MLP model was also used to adjust for sensitivity and specificity of the assay (Uyoga et al, 2021; Gelman et al, 2016).

To estimate the stratum seroprevalence we fitted a Bayesian Multilevel Logistic Regression with poststratification that included sex as a fixed effect, and age and region as random effects (Downes et al, 2018; Uyoga et al, 2021). The model was fit in RJags, a package in R version 3.6.3. Vague or weakly informative priors were used for all parameters and hyperparameters. The Bayesian Multilevel Logistic regression and poststratification model is implemented in two steps:

### STEP 1: MULTILEVEL REGRESSION

The multilevel regression model specifies a linear predictor for the mean J_I_ (or logit transformation of the mean in case of a binary outcome) in some poststratification cell (Downes et al, 2018). We use this model to estimate 238 age-sex-region strata (cells) as decribed earlier. The model specifies that the number of seropositive individuals in age-group (*i*= 1, …, 17), region (*j*= 1, …, 7) and sex (*k* = 1,2) follow a binomial distribution:

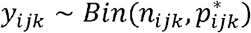

where 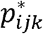 is the observed prevalence and *n*_*ijk*_ is the number tested. For assay test adjustment, the observed prevalence was assumed to depend on the true prevalence as well as the assay sensitivity and specificity (Leeflang et al, 2013; Larremore et al, 2020). The observed prevalence is given by:

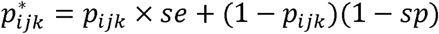

where *se* = *assay sensitivity, sp = assay specificity* and *p*_*ijk*_ is the true prevalence which also depends on sex, age and region of person being tested. The equation below indicate the relationship assumed between true prevalence and the different characteristics in our model:

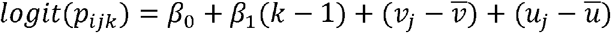

with 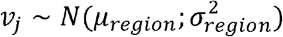 and 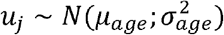 being mean centered random effects for region and age respectively. A similar option would be to fit using the non-centered random effects. For the assay test adjustment, the sensitivity and specificity of Roche were included in the model likelihood assuming a binomial model:

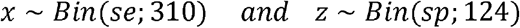

based on the data from a validation study performed by the laboratory. Non-informative priors were used for the intercept and fixed effect coefficients as well as mean components for the random effect model components whilst the half-normal prior was used for age and region variance components.

### STEP 2: POSTSTRATIFICATION

Region, age and sex specific prevalence estimates were then obtained by appropriately weighting the strutum-specific prevalence estimates using data from the 2011 South African Census for Johannesburg. The computation is based on following equation:

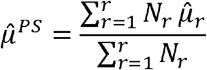

where PS represents postratification, 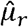 is the estimated seropositive prevalence for poststratification cell *r* and *N*_*r*_ is the size of the *r* poststratification cell in the population. An estimate at any subpopulation level can then be derived by:

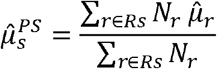

where *Rs*is the subset of all poststratification cells that comprise *s*.

The RJags code is:

~~~
#Seroprevalence using bayesian multilevel regression post weightingting
library(rjags)
library(coda)
### Data
# female = 1, male = 2;
# age categories: 1 = 0-4, 2 = 5-9, 3 = 10-14, 4 = 15-19, 5 = 20-24; 6 = 25-29;
#    7 = 30-34; 8=35-39; 9=40-44; 10=45-49; 11=50-54; 12=55-59;
#    13=60-64; 14=65-69; 15=70-74; 16=75-79; 17=80+
# regions: 1 = Region_A, 2 = Region_B, 3 = Region_C, 4 = Region_D,
# 5 = Region_E, 6 = Region_F, 7 = Region_G
seroprev <- read.csv("seroprevalence_strutum_data.csv")
head(seroprev, 5)
nr <- 7
na <- 17
ns <- 2
y <- array(seroprev$spikepos, dim = c(na, nr, ns))
n <- array(seroprev$n, dim = c(na, nr, ns))
pw <- array(seroprev$pw, dim = c(na, nr, ns))
### Model
model_string_test <- "model{
# Likelihood
for(i in 1:na){
for(j in 1:nr){
for(k in 1:ns){
y[i, j, k] ∼ dbinom(se * p[i, j, k] +
(1 - sp) * (1 - p[i, j, k]),
n[i, j, k])
logit(p[i, j, k]) <- (b0 + b1 * (k - 1)
+ u[i] - mean(u[]) + v[j] - mean(v[]))
}
}
}
# Sensitivity and specificity
x ∼ dbinom(se, 310)
z ∼ dbinom(sp, 124)
# Age effect
tau_a <- 1/pow(sd_a, 2)
for(i in 1:na){
u[i] ∼ dnorm(mu_a, tau_a)
}
# Region effect
tau_r <- 1/pow(sd_r, 2)
for(j in 1:nr){
v[j] ∼ dnorm(mu_r, tau_r)
}
# Priors
b0 ∼ dnorm(0, 1e-04)
b1 ∼ dnorm(0, 1e-04)
se ∼ dbeta(1, 1)
sp ∼ dbeta(1, 1)
# Hyperpriors
sd_a ∼ dnorm(0, 4) T(0,)
sd_r ∼ dnorm(0, 4) T(0,)
mu_a ∼ dnorm(0, 1e-04)
mu_r ∼ dnorm(0, 1e-04)
# Predicted prevalence by age, region and sex
for(i in 1:na){
agecat[i] <- inprod(p[i,1:nr,1:ns], pw[i,1:nr,1:ns])/sum(pw[i,1:nr,1:ns])
}
for(j in 1:nr){
region[j] <- inprod(p[1:na,j,1:ns], pw[1:na,j,1:ns])/sum(pw[1:na,j,1:ns])
}
for(k in 1:ns){
sex[k] <- inprod(p[1:na,1:nr,k], pw[1:na,1:nr,k])/sum(pw[1:na,1:nr,k])
}
JHB <- inprod(p[1:na,1:nr,1:ns], pw[1:na,1:nr,1:ns])
}"
### Compile and update
model_test <- jags.model(textConnection(model_string_test),
  data = list(y = y,
    n = n,
    x = 288,
    z = 122,
    pw = pw,
    na = na,
    nr = nr,
    ns = ns),
  n.chains = 1,
  inits = list(.RNG.name = "base::Wichmann-Hill",
   .RNG.seed = 999))
update(model_test, 1000, progress.bar = "none") # Burn-in period = 1000 samples
samp_test <- coda.samples(model_test,
  variable.names = c("agecat", "region", "sex", "JHB",
    "se", "sp"),
  n.iter = 10000,
  progress.bar = "none")
summary(samp_test)
plot(samp_test)
~~~

## REFERENCES

1. Zhu N, Zhang D, Wang W, Li X, Yang B, Song J, et al. A Novel Coronavirus from Patients with Pneumonia in China, 2019. N Engl J Med. 2020;382:727–33. doi:https://doi.org.10.1056/NEJMoa2001017

2. World Health Organization. Director-General’s opening remarks at the media briefing on COVID-19 - 11 March 2020 [press release].

3. NICD. First case of COVID-19 announced – an update: National Institute for Communicable Diseases; [Available from: https://www.nicd.ac.za/first-case-of-covid-19-announced-an-update/. (xAccessed: 05/01/2021)

4. Abdool Karim SS. The South African Response to the Pandemic. N Engl J Med. 2020;382(24):e95–e. doi:https://doi.org.10.1056/NEJMc2014960

5. NICD. Laboratory confirmed cases of COVID-19 in South Africa: National Institute ForCommunicable Diseases; 2020 [Available from: https://www.nicd.ac.za/wp-content/uploads/2020/07/NICD-Weekly-Epidemiological-Brief_-Week-ending-25-July-2020.pdf. (Accessed: 05/01/2021)

6. WHO. WHO Coronavirus Disease (COVID-19) Dashboard [Available from: https://covid19.who.int/region/afro/country/za (Accessed: 10/01/2021).

7. NICD. Guidelines for case-finding, diagnosis, management and public response (3 July 2020) [Available from: https://www.nicd.ac.za/diseases-a-z-index/covid-19/covid-19-guidelines/ (Accessed: 06/01/2021)

8. Lai CC, Wang JH, Hsueh PR. Population-based seroprevalence surveys of anti-SARS-CoV-2 antibody: An up-to-date review. Int J Infect Dis. 2020;101:314–22. doi:https://doi.org.10.1016/j.ijid.2020.10.011

9. Murhekar MV, Bhatnagar T, Selvaraju S, Rade K, Saravanakumar V, Vivian Thangaraj JW, et al. Prevalence of SARS-CoV-2 infection in India: Findings from the national serosurvey, May-June 2020. Indian J Med Res. 2020;152:48–60. doi:https://doi.org.10.4103/ijmr.IJMR_3290_20

10. Chibwana MG, Jere KC, Kamng’ona R, Mandolo J, Katunga-Phiri V, Tembo D, et al. High SARS-CoV-2 seroprevalence in Health Care Workers but relatively low numbers of deaths in urban Malawi. medRxiv. 2020. doi:https://doi.org.10.1101/2020.07.30.20164970

11. Uyoga S, Adetifa IMO, Karanja HK, Nyagwange J, Tuju J, Wanjiku P, et al. Seroprevalence of anti-SARS-CoV-2 IgG antibodies in Kenyan blood donors. Science. 2021;371:79–82. doi:https://doi.org.10.1126/science.abe1916

12. Hsiao M, Davies MA, Kalk E, Hardie D, Naidoo M C C. SARS-COV-2 seroprevalence in the Cape Town metropolitan sub-districts after the peak of infections: NICD; 2020 [Available from: http://www.nicd.ac.za/wp-content/uploads/2020/06/COVID-19-Special-PublicHealth-Bulletin. (Accessed: 08/02/2021)

13. Gouda HN, Charlson F, Sorsdahl K, Ahmadzada S, Ferrari AJ, Erskine H, et al. Burden of non-communicable diseases in sub-Saharan Africa, 1990-2017: results from the Global Burden of Disease Study 2017. Lancet Glob Health. 2019;7:e1375–e87. doi:https://doi.org.10.1016/S2214-109X(19)30374-2

14. Yang J, Zheng Y, Gou X, Pu K, Chen Z, Guo Q, et al. Prevalence of comorbidities and its effects in patients infected with SARS-CoV-2: a systematic review and meta-analysis. Int J Infect Dis. 2020;94:91–5. doi:https://doi.org.10.1016/j.ijid.2020.03.017

15. Hesse R, vd Westhuizen D, George JA. COVID-19 Related Laboratory Analyte Changes and the Relationship between SARS-CoV-2 and HIV, TB and HbA1c in South Africa. Adv Exp Med Biol 2020. https://doi.org. DOI: 10.13140/RG.2.2.20854.01600/2

16. Mayne ES, Scott L, Semete B, Julsing A, Jugwanth S, Mampeule N, et al. The role of serological testing in the SARS-CoV-2 outbreak. S Afr Med J. 2020;110:842–5.

17. Sykes W, Mhlanga L, Swanevelder R, Glatt TN, Grebe E, Coleman C, et al. Prevalence of anti-SARS-CoV-2 antibodies among blood donors in Northern Cape, KwaZulu-Natal, Eastern Cape, and Free State provinces of South Africa in January 2021. doi:https://doi.org.0.21203/rs.3.rs-233375/v1

18. NICD. Dominance of the SARS-CoV-2 501Y.V2 lineage in Gauteng 2021 [Available from: https://www.nicd.ac.za/wp-content/uploads/2021/01/Dominance-of-the-SARS-CoV-2-501Y.V2-lineage-in-Gauteng-South-Africa-1.pdf. (Accessed: 02/03/2021)

19. Kufa-Chakezha T, Jassat W, Walaza S, Erasmus L, von Gottberg A, Cohen C. Epidemiology and clinical characteristics of laboratory confirmed COVID-19 among individuals aged ≤19 years, South Africa, 1 March 2020 – 2 January 2021: NICD; 2021 [updated 2021. Available from: http://www.nicd.ac.za/WP-content/uploads/2021/01.COVID-19-SPECIAL-PUBLICHEALTH-SURVEILLANCE-BULLETIN-Volume-7. (Accessed: 02/03/2021)

20. Hippich M, Holthaus L, Assfalg R, Zapardiel-Gonzalo J, Kapfelsperger H, Heigermoser M, et al. A Public Health Antibody Screening Indicates a 6-Fold Higher SARS-CoV-2 Exposure Rate than Reported Cases in Children. Med (N Y). 2021;2:149-63.e4. doi:https://doi.org.10.1016/j.medj.2020.10.003

21. Sahasranaman A, Jensen HJ. Spread of COVID-19 in urban neighbourhoods and slums of the developing world. J R Soc Interface. 2021;18:20200599-. doi:https://doi.org.10.1098/rsif.2020.0599

22. Singh AK, Gupta R, Ghosh A, Misra A. Diabetes in COVID-19: Prevalence, pathophysiology, prognosis and practical considerations. Diabetes Metab Syndr. 2020;14:303–10. doi:https://doi.org.10.1016/j.dsx.2020.04.004

23. Mutyambizi C, Booysen F, Stokes A, Pavlova M, Groot W. Lifestyle and socio-economic inequalities in diabetes prevalence in South Africa: A decomposition analysis. PLoS One. 2019;14:e0211208–e. doi:https://doi.org.10.1371/journal.pone.0211208

24. Suhandynata RT, Hoffman MA, Huang D, Tran JT, Kelner MJ, Reed SL, et al. Commercial Serology Assays Predict Neutralization Activity Against SARS-CoV-2. Clin Chem. 2020. doi:https://doi.org.10.1093/clinchem/hvaa262

## References

1. Gelman, A., Lax, J., Phillips, J., Gabry, J., & Trangucci, R. (2016). Using multilevel regression and poststratification to estimate dynamic public opinion. Unpublished manuscript, Columbia University, 2.

2. Uyoga, S., Adetifa, I. M., Karanja, H. K., Nyagwange, J., Tuju, J., Wanjiku, P., … & Warimwe, G. M. (2021). Seroprevalence of anti–SARS-CoV-2 IgG antibodies in Kenyan blood donors. Science, 371(6524), 79–82.

3. Downes, M., Gurrin, L. C., English, D. R., Pirkis, J., Currier, D., Spittal, M. J., & Carlin, J. B. (2018). Multilevel regression and poststratification: A modeling approach to estimating population quantities from highly selected survey samples. American journal of epidemiology, 187(8), 1780–1790.

4. Leeflang, M. M., Rutjes, A. W., Reitsma, J. B., Hooft, L., & Bossuyt, P. M. (2013). Variation of a test’s sensitivity and specificity with disease prevalence. CMAJ : Canadian Medical Association journal = journal de l’Association medicale canadienne, 185(11), E537–E544. https://doi.org/10.1503/cmaj.121286

5. Larremore, Daniel B., Bailey K. Fosdick, Kate M. Bubar, Sam Zhang, Stephen M. Kissler, et al. Estimating SARS-CoV-2 seroprevalence and epidemiological parameters with uncertainty from serological surveys (2020).

